# A Structural Equation Model Predicts Chronic Wound Healing Time Using Patient Characteristics and Wound Microbiome Composition

**DOI:** 10.1101/2025.01.23.25320984

**Authors:** Jacob Ancira, Rebecca Gabrilska, Craig Tipton, Clint Miller, Zachary Stickley, Khalid Omeir, Catherine Wakeman, Todd Little, Joseph Wolcott, Caleb D. Philips

## Abstract

Wound etiology, host characteristics, and the wound microbiome contribute to chronic wound development. Yet, there is little accounting for the relative importance of these factors to predict wound healing. Here, a structural equation model was developed to provide such an explanatory and predictive framework. Chronic wounds from 565 patients treated at a clinic practicing biofilm-based wound care were included. Patient information included DNA sequencing-based wound microbiome clinical reports corresponding to initial clinical visit. Wound microbiome data was integrated into the SEM as a latent variable using a pre-modeling parcel optimization routine presented herein for the first time (available as R library *parcelR*). A microbiome latent construct associated with improved healing was validated, and the final SEM included this latent construct plus three species associated with diminished healing (*Anaerococcus vaginalis, Finegoldia magna, Pseudomonas aeruginosa*), as well as smoking, wound volume, slough, exudate, edema, percent granulation, and wound etiology This model explained 46% of variation in healing time with the microbiome contributing the largest proportion of variance explained. Model validity was confirmed with an independent cohort (n = 79) through which ∼60% of variation in healing time was predicted. This model can serve as foundation for development of a predictive tool that may have clinical utility.

## 1 INTRODUCTION

Chronic wounds are commonly defined as wounds that fail to show signs of healing within three weeks^1^ and are a tremendous burden on the healthcare system with the elderly and diabetics being predominantly affected.^2^ The cost of chronic wounds in the United States has been estimated in the billions, with around 8.2 million people suffering from chronic wounds annually.^3^ Chronicity has a multifaceted basis, and its emergence and maintenance is widely recognized to be promoted by the colonization of microbes,^4,5^ and the resulting communities are commonly referred to as the wound microbiome. The relationship between chronic wound formation, persistence, and wound microbiome is particularly important because differences in wound microbiomes are thought to be consequential to healing.^6^ The recognized role of microbes in wound chronicity is the basis for biofilm-based wound care approaches aiming to reduce microbial load and biofilm in chronic wounds to improve healing times.^7^

Several studies have been conducted to understand wound microbiome composition. For example, chronic wound microbiomes have been reported as compositionally distinct from that of adjacent healthy skin^8^ from which they are thought to be frequently colonized.^9^ A study of about 3,000 chronic wound samples from patients in the south-central USA found most wound microbiomes to be polymicrobial with the five most common bacterial species being *Staphylococcus aureus*, *Staphylococcus epidermidis*, *Pseudomonas aeruginosa*, *Finegoldia magna*, *Stenotrophomonas maltophilia*.^4^ Other work often reports a partially overlapping list of most common taxa (e.g., Townsend et al. 2023).^10^ It is thought that the colonization of microbes into chronic wounds is a partially deterministic process. For example, diabetes and obesity can cause decreased blood flow to wounds, resulting in environmental selection for anaerobic bacteria.^11^ Other work indicates that differences in patients’ genetics are predictive about the types of chronic infections people develop, and infecting communities may further be shaped by microbial species’ interactions.^12^

Previous research has investigated how wound microbiomes explain differences in healing. For example, Losche et al. (2017) reported how longitudinal trends in wound microbiome composition relate to healing, with compositional instability associated with better outcomes.^6^ Other recent work integrating wound properties, proteomics and microbiome into a multivariate discriminant analysis suggested distinct characteristics of healed versus ongoing wounds.^10^ Such work is motivated by the idea that better understanding the microbiome’s role in chronicity and healing will yield mechanistic insights and could be used to improve wound care. In other words, being able to confidently estimate a patient’s healing time based on their characteristics could result in a better understanding of the basis for responsiveness to treatment and may eventually guide therapy.

One of the challenges of characterizing microbiome effects on healing is how to account for the large number of combinations of species’ abundances and their interactions and how those relate to healing differences. Relating individual or multiple species to healing has been done by directly comparing their incidence or abundance to healing times or endpoints^12,13^ and include using species relative abundances in model feature selection.^10^ Reducing multivariate microbiome data to eigenvectors of an ordination, then using those axes as predictor variables has also been demonstrated.^14^ Other potential approaches include relating healing to wound microbiome alpha (within site diversity, e.g., richness) or beta (compositional similarities among wound microbiomes) diversity; however, these approaches are not well-suited to investigate species effects and the summarization of a sample’s multi-species abundances as single values results in information loss.

Structural Equation Modeling (SEM) is a general linear framework for modeling latent variables and has the potential to reveal more about the relationship between microbiome composition and host characteristics, specifically healing in the case of chronic wounds. This may be accomplished by modeling wound microbiomes as a latent construct estimated as the linear solution of groups of species’ abundances correlating with healing time. Such microbiome latent variables may contain contributions of individual species as well as emergent community effects on healing. Recognizing the important knowledge gap on understanding the role of microbes and patient variables on healing outcomes, as well as the potential long-term clinical utility of such knowledge, the goal of this study was to develop and validate a model for healing time.

## 2 MATERIALS & METHODS

### 2.1 Sample and Patient Data Collection

Deidentified patient data included in this study are from patients that received treatment at the Southwest Regional Wound Care Center (Lubbock, TX) for chronic wound infections (IRB protocol IRB2016-1073). The wound care protocol for patients involved wound microbiome compositional profiling at initial clinic visit with subsequent follow-up visits until healing, which was determined by the physician upon complete closure of the wound. The full treatment protocol is as follows, upon admission to the clinic for care, the wound is examined for various measurements such as the amount of edema, slough, and accumulation of fluid. If debridement is needed, the following procedure is implemented. 1) The wound is prepared by applying 2% Lidocaine Gel to the wound. 2) The wound is cleansed with normal saline, and a clean field is provided surrounding the wound. 3) By sharp excisional debridement using iris scissors, pickups, and a curette, all devitalized tissue, biofilm/slough, and wound margins are removed to expose viable, normal, bleeding tissue throughout the wound bed. 4) An Anti-Psuedomonas Gel (APG) made using Distilled water, Gentamicin, Benzalkonium Chloride, citric acid, Clindamycin, lidocaine, and Methylcellulose 4000cps is applied to all wounds with various dressing dependent on the amount of exudate observed. Finally, a treatment plan is created depending on the type of wound and various characteristics observed. For example, compression is applied to wounds with edema, lymphedema, and/or venous leg ulcers or offloading in the case of diabetic foot ulcers and decubitus ulcers. Treatment varies case to case but is conducted in accordance with Wound Healing Society guidelines.

Wound microbiome compositional profiles were obtained by sending homogenized wound debridement, obtained from step 3 of the previously described method, to MicroGenDX (Lubbock, TX), which is a CAP and CLIA certified high-complexity diagnostic laboratory. The molecular and bioinformatic methods employed at MicroGenDX for bacterial V1-V2 16s rRNA gene profiling have been described in detail elsewhere^15^ and bacterial species relative abundances from resulting clinical reports for each patient were tabulated for analysis. Patient metadata associated with each sample were retrieved from the electronic medical records and are described below. Wounds were photographed and measured at the initial visit to determine wound size.

### 2.2 Structural Equation Modeling

#### 2.2.1 Microbiome Parceling

Structural Equation modeling (SEM) is an analytical approach to modeling and estimating latent variables. What makes a variable latent is its inability to be accurately measured with a single discrete or continuous variable measurement. Latent constructs in an SEM are estimated as the weighted best linear solution of indicator variables given the model, and indicator variables are those which can be directly measured and are thought to describe the latent construct. Here, relative abundances of species served as the indicator variables for a microbiome latent construct. However, model fit in SEM constrains latent variable specification to ideally three indicators, but the number of species potentially important to inform a latent microbiome construct relating to healing may be larger.

A common solution to the problem of many indicators is item parceling, through which variables are grouped and averaged, and the resulting parcels serve as the indicators.^16^ However, item parceling has not previously been adapted to metagenomic data and is described here for the first time. The optimal parceling scheme amongst several species for the microbiome data was identified by 1) randomly assigning species to one of three parcels, 2) averaging the log transformed relative abundances of species within each parcel, 3) recording inter-parcel correlations and variances, and 4) recording parcel-healing time correlations and variances. This process of random parcel assignment, averaging, and correlation and variance recording was repeated 1,000 times, and comparison amongst the 1,000 parceling schemes was then performed to identify the best scheme.

Comparison was made using a multi-objective optimization approach, in which the objective was to jointly maximize inter-parcel and parcel-healing time correlative relationships. This was achieved by a) dividing the summed inter-parcel correlation by its variance followed by proportion of maximum (POM) transformation (referred to as the scaled parcel correlative relationship), b) dividing the summed parcel-healing time correlation by its variance followed by POM transformation (referred to as the scaled parcel explanatory strength), and c) identifying the best parceling scheme as that with the smallest Euclidian distance from the theoretical best parceling scheme which would score 1 for both metrics described in steps a and b. This iterative approach of parcel identification was performed on subsets of the data by creating species subsets based on positive and negative Pearson correlations with healing time in increments of 0.1 in either direction. The best candidate positively and negatively associated microbiome latent variables were identified based on having highest R^2^ and a significant p-value from confirmatory factor analysis. The validity of resulting latent constructs was assessed by comparison to null distributions of R^2^ values obtained by randomly assigning healing times to patients and iterating the above-described routine at the corresponding correlation threshold 1000 times (Figure 1). The procedure described above for optimal parcel discovery is made available via the R library, *parcelR* available at https://github.com/jakcira.

**Figure 1.**
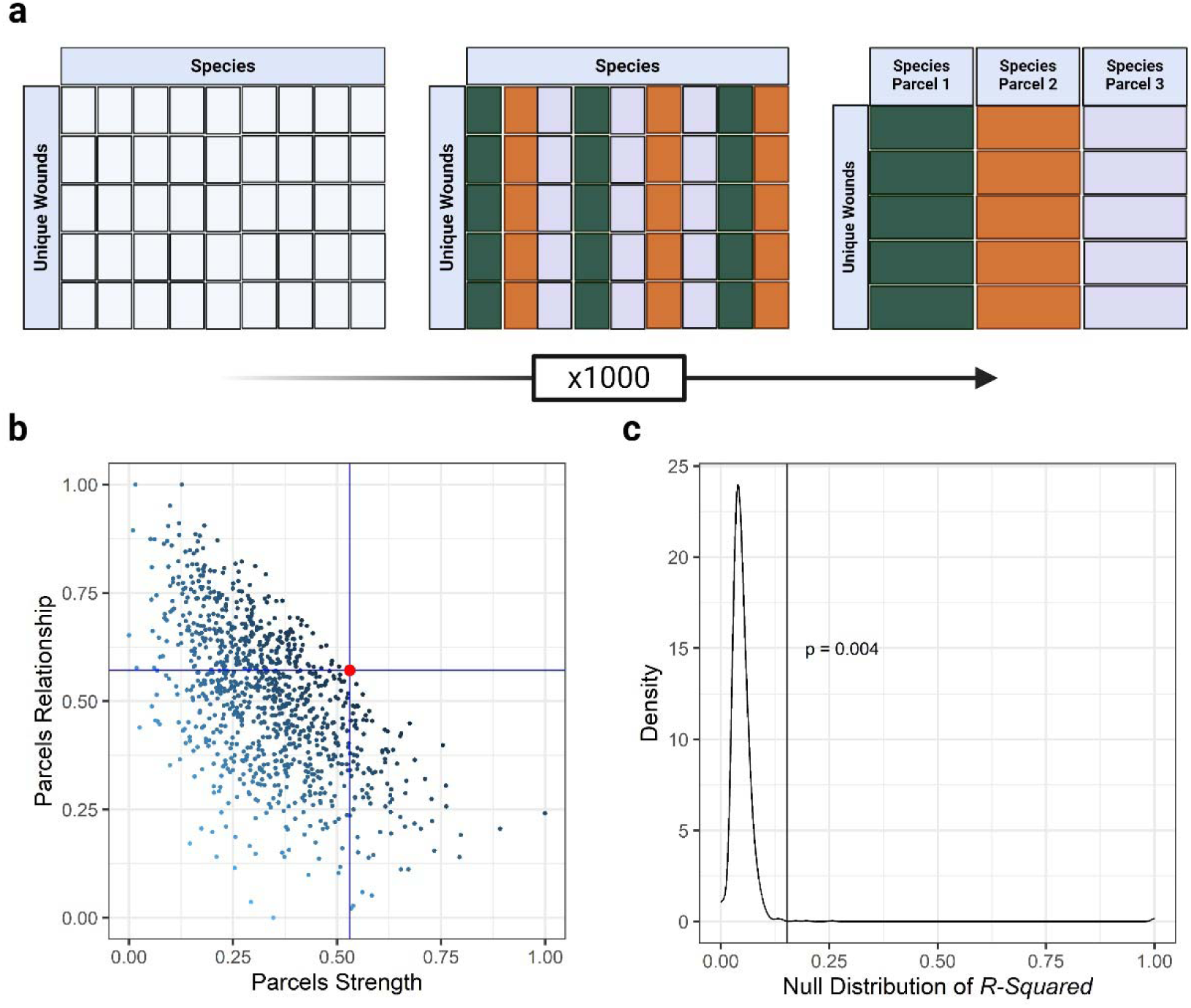
Illustration of the parcel optimization routine. A) Beginning with a log transformed table of patients by species, each species is randomly assigned to one of three parcels. The abundances of species assigned to each parcel are averaged, and their correlations and variance with each other and the outcome of interest (healing time) is recorded. B) The process described in A is iterated many times (1000, herein), and the best parceling scheme (denoted by the red dot) is identified as that which jointly maximizes the relationships amongst parcels (scaled parcel correlative relationship) and to the outcome (scaled parcel explanatory strength. C) The validity of the optimal parceling scheme is tested by comparison of the observed R^2^ for healing time (vertical line) to a null distribution of R^2^ values based on 1,000 iterations of the procedure described in panels A and B, but by first randomly pairing healing time and microbiomes.

#### 2.2.2 Model Variables

To further consider species associated with healing time but not included in the parcels, species were filtered by correlation p-value < 0.05 and incidence of at least 5% in the patient sample. Other candidate predictor variables describing patients and their wounds included in model evaluation were race, sex, age, BMI, wound volume, smoker status, granulation, slough, exudate, edema, erythema, wound type classification variables, pain, wound location, and comorbidities. Granulation, slough, exudate, edema, erythema, and pain were scored according to the proposed modified TIME-H comorbidity score.^17^ These scores were reversed (2 = Severe, 1 = medium, 0 = mild/none) to be in a logical order with the expectations of relationship to healing time. Additionally, to score comorbidities more accurately, the Charlson Comorbidity Index was applied by first translating diagnosis to ICD 10 codes then calculating the overall score for each patient.^18,19^ Wound volume was modeled as a latent variable manifest by width, depth, and length. To detect any effect of wound type independent of microbiome composition and wound volume, separate latent constructs were created for each wound type that were independent of microbiome and wound volume latent variables.

#### 2.2.3 Model Identification

Model identification used backward selection based first on p-value followed by significance testing for a difference in model χ^2^, and variables were iteratively removed until removing additional variables results in significantly worse model fit. After a sufficient pruned model was reached, the manifest variables were modeled as phantom latent variables.^20^ These latent variables were created using a single indicator manifest variable with a 100% loading onto the phantom latent. This was done for every manifest variable to understand the proportion of the overall *R^2^* accounted for by each variable.

The acceptability of the final model was jointly determined by the four main fit measures p-value, Comparative Fit Index (CFI), Tucker-Lewis Index (TLI), and Root Mean Squared error of approximation (RMSEA). The latter three metrics of evaluating model fit are all derived from the use of the χ^2^ and degrees of freedom. The χ^2^ alone was not used as an indicator of model fit due to its sensitivity to moderately larger data sets. Proposed thresholds for interpreting model fit using RMSEA used here include good/close fit (0.05 – 0.02), acceptable fit (0.08 - 0.05) and poor/mediocre fit (> 0.08).^21,22^ For CFI, above 0.9 is considered an acceptable model fit, and 0.95 – 0.99 being a very good fit. TLI is judged on the same scale and is highly correlated with the CFI but tends to be slightly lower than the CFI due to a higher penalty for model complexity.

### 2.3 Species Importance

The importance of individual species to microbiome parcels was estimated by a leave-one-out analysis. For this analysis the difference in *R^2^* value between the final SEM and SEMs in which single species were iteratively removed from the microbiome parcels were recorded. Also, to explore if parceled species’ contribution to healing were solely due to their direct effects, or also due to species interactions, the species with the largest Δ*R^2^* from the leave-one-out analysis was used in a new model with the microbiome latent replaced by this species relative abundance, and the *R^2^* difference of these models were compared. Binomial testing was used to assess if species that were incorporated into parcels were non-random subsets with respect to the frequencies of gram staining, aerotolerance, species, and genera frequencies in the total dataset. Species cooccurrences, and significance of cooccurrences were calculated and chi-squared testing was used to see if cooccurrence rates differed depending on if species were included in the microbiome latent construct.^23^ A t-test was used to assess if the distribution of Pearson correlations significantly differed among parceled vs non-parceled species.

### 2.4 Model Validation

Because using principal component analysis (PCA) as a means of microbiome variable reduction to serve as input into SEM has been previously reported,^14^ the final model was compared to a model fit using PCA eigenvectors in the place of the identified microbiome parcels. The PCA eigen vectors were identified (*princomp* function*, stats* R package) and selected for inclusion based on a broken stick regression (*screeplot* function*, stats* R package). Three eigen vectors were included in the saturated model as manifest variables followed by the previously described backwards selection process.

An independent cohort of 79 unique wounds were used to evaluate the performance of the final model in predicting wound duration. This was accomplished by multiplying observed data by their corresponding model regression coefficients (β) then summed to obtain the predicted duration. The predicted duration was modeled as a function of the observed duration and the *R^2^* and *p-value* from this regression were the metrics of performance.

## 3 RESULTS

### 3.1 Data Characterization

The final dataset included 565 patients that were classified by clinicians into four wound types described as: atypical wound, diabetic foot ulcer, decubitus ulcer, venous leg ulcer, and non-healing surgical wound (Table 1, Supplementary Table 1). The atypical wound category is a general category to which wounds without clear etiology were assigned. Among the 452 total bacterial species reported in at least one patient’s clinical report there were 110 aerobes and 342 that were either anaerobes or facultative anaerobes. Species were further described by staining with 240 Gram positive and 212 Gram negative. The top five most common genera were *Corynebacterium*, *Staphylococcus*, *Streptococcus*, *Bacteroides*, and *Prevotella*. Each of these genera consisted of 33, 21,18,18, and 16 unique species, respectively and consisted of some of the most abundant species. The top five most detected species were *Staphylococcus aureus*, *Staphylococcus epidermidis*, *Finegoldia magna*, *Anaerococcus vaginalis*, and *Streptococcus agalactiae*. Most (87.9%) wounds were reported as polymicrobial. Refer to Supplementary Table 2 for a list of all non-polymicrobial wounds and what species was observed.

**Table 1.** Patient demographics and wound characteristics. Cell values are either means with parenthetical standard deviations, or n with parenthetical percent of total n.

| Characteristic | N = 565 | Characteristic | N = 565 |
| --- | --- | --- | --- |
| Duration (Days) | 396 (519) | Slough | 23 (16) |
| Exudate |  | Percent Granulation | 64 (47) |
| Copious | 11 (1.9%) | Wound Type |  |
| Mild | 17 (3.0%) | Decubitus Ulcer | 42 (7.4%) |
| Minimal | 197 (35%) | Diabetic Foot Ulcer | 105 (19%) |
| Moderate | 150 (27%) | Atypical Wound | 385 (68%) |
| None | 123 (22%) | Venous Leg Ulcer | 33 (5.8%) |
| Severe | 67 (12%) | Race |  |
| Erythema |  | Black | 45 (8.0%) |
| Mild | 308 (55%) | Hispanic | 167 (30%) |
| Moderate | 123 (22%) | Other | 1 (0.2%) |
| None | 117 (21%) | White | 352 (62%) |
| Severe | 17 (3.0%) | Smoker | 100 (18%) |
| Edema |  | Age | 62 (14) |
| Mild | 2 (0.4%) | Height (in) | 67.3 (9.4) |
| Minimal | 261 (46%) | Weight (lbs) | 212 (64) |
| Moderate | 155 (27%) | Wound Depth (cm) | 0.22 (0.33) |
| None | 122 (22%) | Wound Width (cm) | 0.55 (1.11) |
| Severe | 25 (4.4%) | Wound Length (cm) | 0.58 (1.16) |

### 3.2 SEM Model Parameter Estimates

The microbiome parceling procedure resulted in two candidate microbiome latent constructs, one each associated with delayed and faster healing times. However, only the latent construct associated with faster healing times could be validated (Figure 1C, p = 0.004). This latent construct was formed from parcels comprised of 66 species exhibiting at least a −0.2 Pearson correlation with healing time (The full list of the 66 species is reported in Supplementary Table 3). Included species had an average incidence rate of 14.3 ± 12.95 patients. Sixty percent (18 species) of the 30 most common species based on study-wide relative abundance were included in this latent variable.

The final SEM model after backwards selection and Δχ^2^ testing consisted of the healing-associated microbiome latent variable, wound volume latent variable, edema, exudate, percent granulation, slough, smoker status, venous leg ulcer, as well as *Pseudomonas aeruginosa*, *Finegoldia magna*, and *Anaerococcus vaginalis* (Figure 2). Good to very good overall model fit was indicated by CFI = 0.950, TLI = 0.937, and RMSEA = 0.034(0.029 – 0.038).

**Figure 2:**
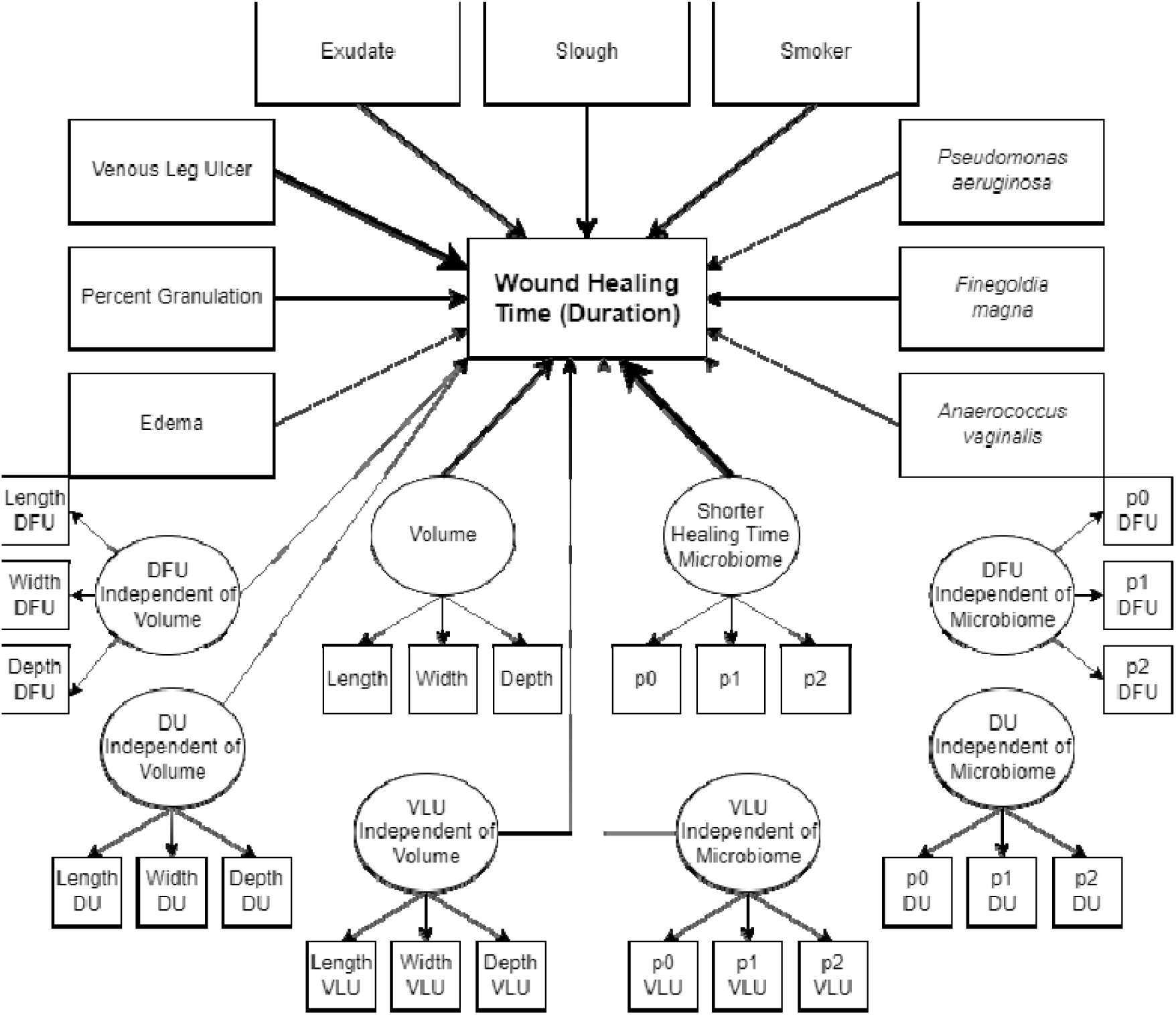
Structural equation model to predict wound healing time: Latent variables are denoted with ovals which are manifested by their corresponding indicators. Parcels are represented by p0, p1, and p2. Residuals, variances, and covariances are reported in Supplementary Table 1. Model parameters are reported in Table 1. Regression arrows are scaled according to the proportion of contribution to the overall R^2^.

The overall *R^2^* of the final model was 0.46, with microbiome accounting for the largest proportion of variance explained at 39.98% with a beta coefficient of −0.632. The next most influential variables were ‘venous leg ulcer’, accounting for 35.82% of variance with a coefficient of −0.598, slough accounting for 7.384% of variance with a coefficient of −0.272, smoking status, accounting for 6.94% of variance with a coefficient of 0.263, percent granulation, accounting for 4.71% of variance with a coefficient of −0.217, wound volume accounting for 3.99% of variance with a coefficient of 0.199, and exudate accounting for 4.84% of variance with a coefficient of 0.220 (Table 2). The remaining variables each explained 2% or less of variance in healing time. Several significant covariances were also recovered among wound environment variables. For example, positive standardized covariances were observed in all pairwise combinations among edema, exudate, and slough. Another set of positive standardized covariances were observed in comparisons of wound volume to *Anaerococcus vaginalis*, *Psuedomonas aeruginosa*, edema, exudate, and slough. Conversely, negative standardized covariances were observed in comparisons of granulation to edema, exudate, slough and venous leg ulcer, among others.

**Table 2:** Regressions of the modeled variables onto wound healing time including variance explained. The proportion of R^2^ (%) refers to the percentage of the full models R^2^ (0.462) contributed by each variable.

| **Variable Regressed onto Duration** | **Regression ($\beta$)** | **Proportion of $R^2$ (%)** |
| --- | --- | --- |
| Microbiome | -0.6323 | 39.98025 |
| Volume | 0.199725 | 3.988998 |
| Smoker | 0.263386 | 6.937194 |
| Slough | -0.27175 | 7.384651 |
| Exudate | 0.220016 | 4.840685 |
| VLU | -0.59849 | 35.81914 |
| Percent Granulated | -0.21704 | 4.710803 |
| Edema | -0.0163 | 0.026568 |
| *Anaerococcus vaginalis* | 0.035428 | 0.125514 |
| *Finegoldia magna* | 0.029659 | 0.087965 |
| *Pseudomonas aeruginosa* | -0.00288 | 0.000832 |
| VDFU | 0.050955 | 0.259644 |
| VVLU | 0.023105 | 0.053384 |
| VDU | -0.06214 | 0.386191 |
| pDFU | 0.057325 | 0.32862 |
| pDU | 0.048017 | 0.230564 |
| PVLU | -0.00022 | 4.94E-06 |

### 3.3 Species Correlations

To gain insights into any microbial characteristics promoting the inclusion of species into the latent variable beyond their positive association with healing, binomial tests were implemented to assess if the frequency of aerotolerance classifications, Gram staining, and genera occurred at a frequency among parceled species different than expected based on study wide frequencies. In no cases were the observed frequencies within parcels significantly different that study wide (p >0.05).

To estimate the impact of each parceled species on the overall model, a leave one out approach was implemented in which each species was iteratively removed from the parceling scheme and the *R^2^* of the resulting model was recorded. The top three species that affected the *R^2^* were *Staphylococcus aureus*, *Corynebacterium striatum*, and *Proteus mirabilis* with Δ*R^2^* of 0.234, 0.071, and 0.064, respectively (Table 3).

**Table 3:**
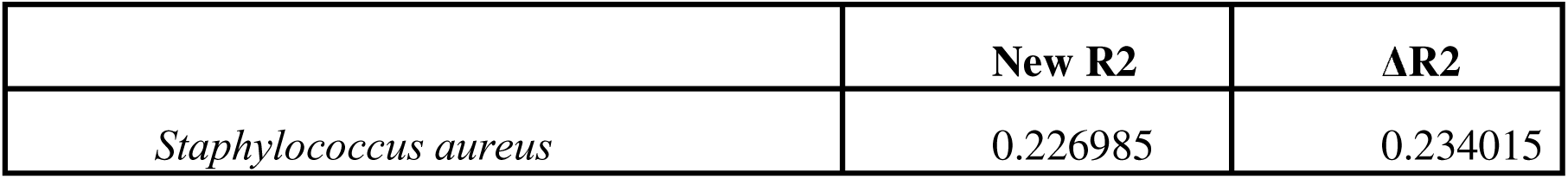
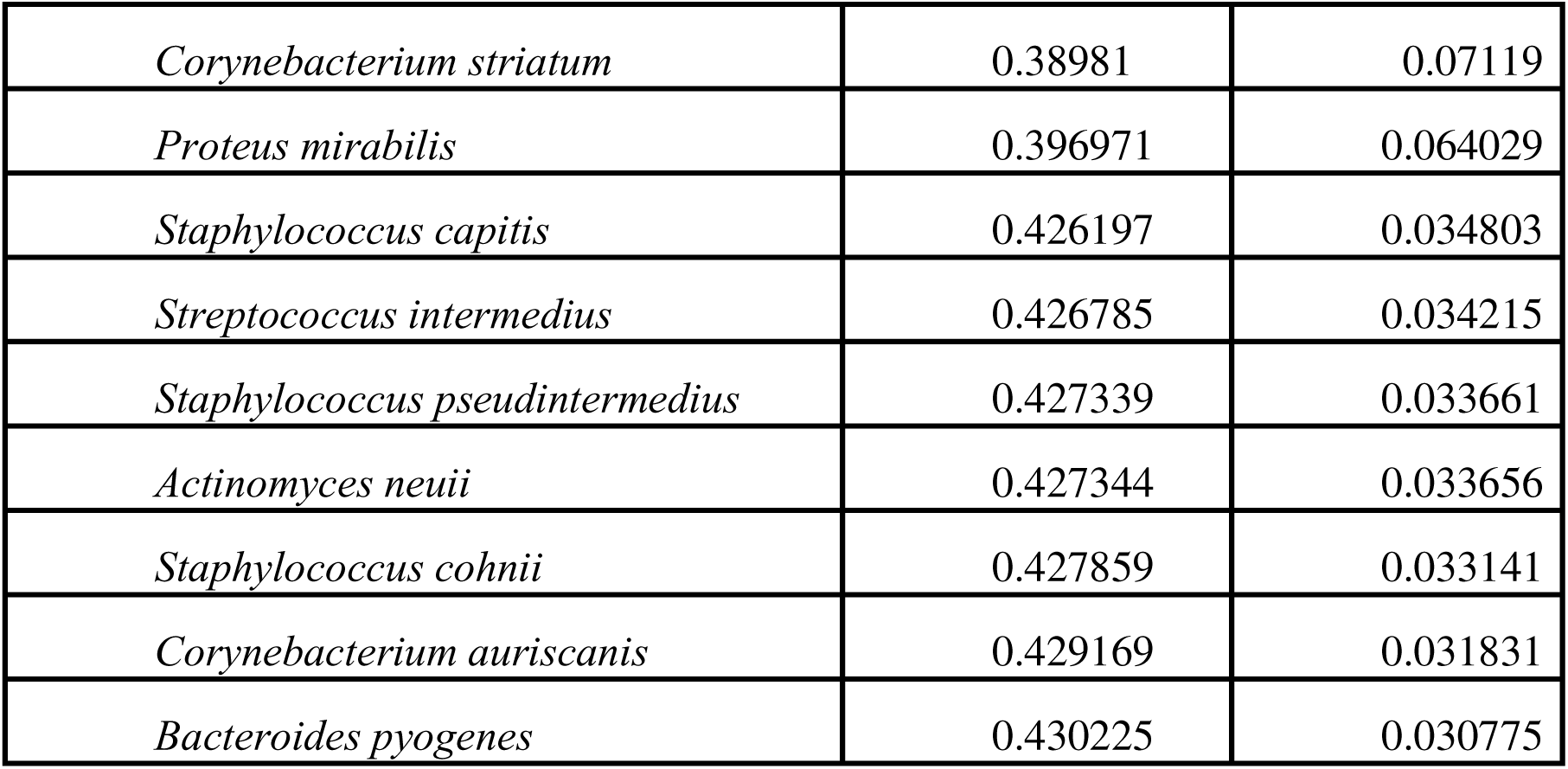
The top 10 most influential species on the overall R^2^ when removed from the microbiome latent construct. The first column shows the R^2^ after removal from the construct, and the last column shows the difference in R^2^ relative to the full model.

Given the outsized role of some species on the *R^2^* of the model, an additional comparison was made to assess the extent such influence was due to species’ direct effect on healing or also contributed by species interactions. For this, the microbiome latent variable was replaced with the log transformed relative abundance values of *S. aureus*, which was selected for exploration based on it having the largest Δ*R^2^*. Whereas the full model explained 46.2%, the resulting *S. aureus* model explained only 5.3% of variance of wound healing time. Moreover, the exclusion of *S. aureus* from the microbiome latent resulted in 9.1% of variance explained.

Finally, assessment of the relative performance of the parceling approach as compared to PCA eigenvectors was made. For this, three PCA axes identified as significant through broken stick analysis were added to the overall SEM model as manifest variables and the microbiome latent variable was removed. Using the same backward selection routine described above for model identification all 3 axes were removed and resulted in a pruned model with *R^2^* = 0.093.

### 3.4 Model Validation

An independent cohort of 79 new patients were used to assess model validity and performance. Here, the latent variable for wound microbiome was estimated using the same species identified in the original model and by constraining the loadings of the parcels onto the latent construct (*lavPredict* function, *lavaan* R package). Similar constraint was employed for the wound volume latent variable. The latent and manifest variables for this cohort were multiplied by the corresponding beta coefficients of the final model, and the summation of these products was the predicted healing time in days. A regression of predicted healing time as a function of observed healing time resulted in an *R^2^* of 0.6 (Figure 3, p < 0.001). On average, the model fit predicted duration 3.02 ± 13.27 weeks lower than the observed duration.

**Figure 3:**
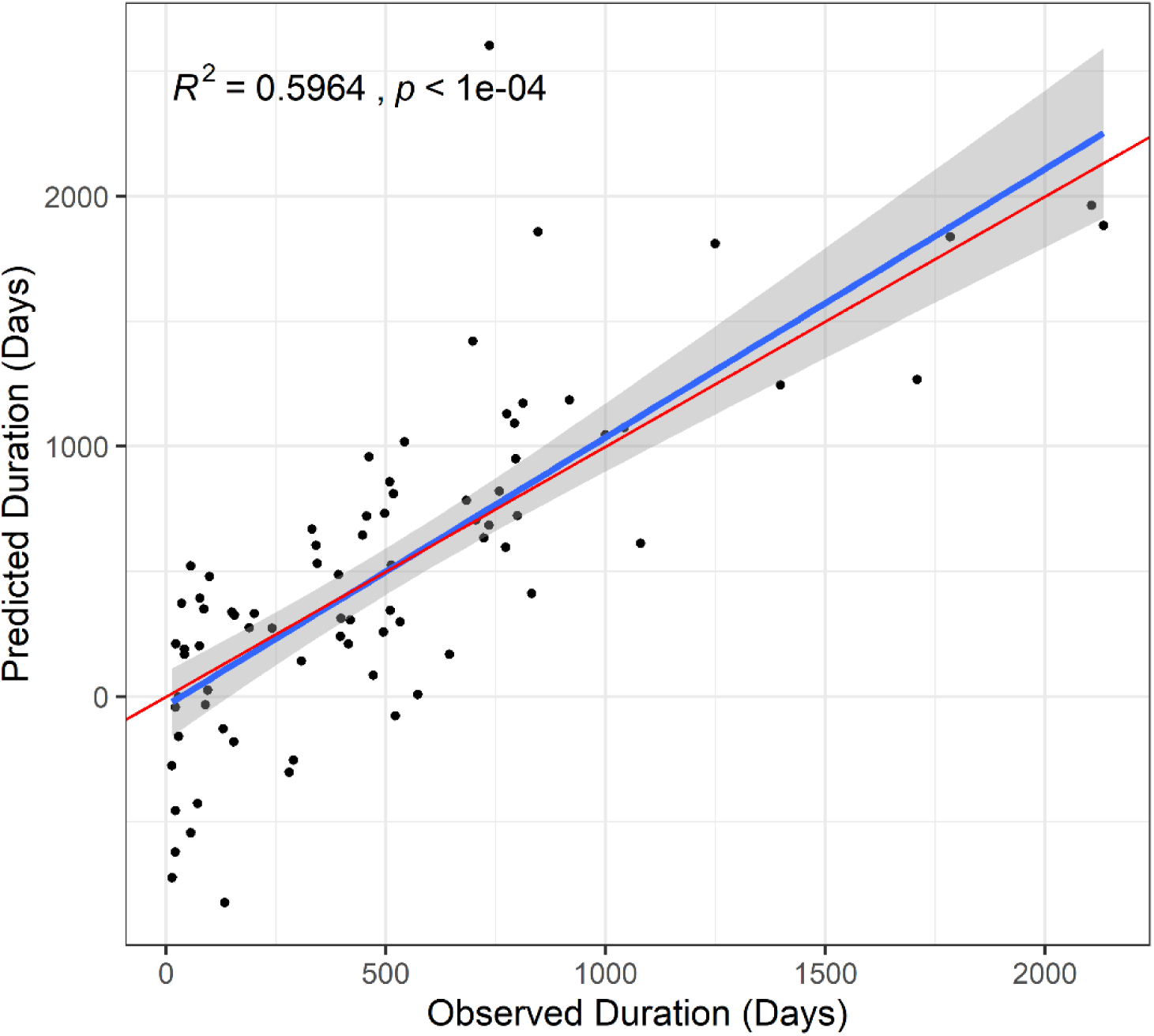
An independent cohort of 79 new wounds was used to validate model performance. For these wounds predicted healing time was calculated using the model parameters calculated in the original cohort, then regressed onto observed healing time to obtain R^2^ as a measure of model performance. The red line illustrates a slope of 1 and is provided for comparison to the predicted slope.

## 4 DISCUSSION

Many aspects of chronic wound persistence and physiology are routinely considered from both research and clinical perspectives as contributors to healing. Leveraging SEM, this study quantifies variable causal influence and covariances as predictors of chronic wound healing. A major result from the final model was that measured patient and clinical parameters explained about half of the variation among patients in their healing time in the initial model (*R^2^* = 0.46), and the model performed even better in a follow-up cohort (*R^2^* = 0.60, Figure 3). Among significantly predicting variables, wound microbiome composition accounted for the most variation in healing time. Generally, this work is unique and the first to utilize the application of SEM to understand factors shaping chronic wound healing. Also, whereas a few other microbiome studies have used SEM, those studies have either relied on manifest variables such as specific microbes^24^ or used eigenvectors as an approach to microbiome variable reduction.^14^ The parceling approach presented herein integrating microbiome compositional information into a latent SEM framework is novel and provides an opportunity for increased explanatory power.

Whereas species varied in their contribution to the explanatory strength of the model (Table 3), comparative analysis support that these contributions were not solely a feature of their direct effect on healing but that their interactions with other species also contributed to the latent variable’s explanation of healing time variation. This was supported by the observation that comparison of the final model to a model in which the microbiome latent variable was replaced by *S. aureus* relative abundance (the species with the strongest Δ*R^2^* when excluded from the latent) had a 40.8% reduction in overall model explanation. Additionally, PCA was used instead of parceling to compare methods. Using PCA axes instead of parcels in the SEM resulted in significantly worse explanation with a 36.9% reduction in the overall model explanation. This contrast illustrates the utility of species parceling and latent variable estimation beyond using species relative abundances or eigenvectors as manifest variables. It is anticipated that this approach will be generally useful for relating microbiome data to continuous variables.

Through the final model’s RMSEA (0.034), CFI (0.95), and TLI (0.93), a good to very good model fit was achieved. The fit of this model is reflected in its explanatory strength on healing time (46%) as well as the large number of significant covariances among variables, all of which vary in directions that are clinically logical. For example, identifying as a smoker significantly associated with longer healing times. Not only does smoking affect the amount of oxygen hemoglobin can carry and reduce oxygen supply to wounds, but it also causes narrowing of blood vessels which slows the rate of blood and nutrients supplied to wounds.^25^ Also, measures of chronic wound physiology associated with chronicity including wound volume, exudate, edema, and slough^26^ all exhibited significant positive pairwise covariances amongst each other as well as to smoking, but all also had significant negative covariances with percent granulation (a marker of healing) as well as the latent microbiome construct predicting faster healing times. Such covariances parallel many interactions previously reported in isolation and serve to validate the model of the wound environment.

Venous leg ulcer classification (VLU) was significantly associated with faster healing times. The inclusion of VLUs in the full model, opposed to the other wound types, is concordant with previous findings due to VLUs most commonly being associated with decreased healing times. Fife (2018) showed healing rates by the 12-week mark was the largest among VLUs (44.1%) when compared to DFUs (30.5%) and Pressure Ulcers (29.6%).^27^ Additionally, <u>Läuchli</u> et al. (2013) reported that among leg ulcers, VLUs had higher healing rates after 6 months (63%) when compared to mixed arterial-venous ulcers (30%) and hypertensive ischemic leg ulcers (35%).^28^ These previous findings are also supported in the current data as VLUs were found to significantly account for faster healing wounds when compared to other wound types (p = 0.004).

The final validated microbiome latent variable associated with faster healing times was formed by a diverse set of bacterial species including less common as well as 18 of the top 30 most abundant bacterial species. There is potential that some of the parceled species are beneficial, such as *Alcaligenes faecalis,* which we show correlated with faster healing and has been shown to improve wound healing within a murine model.^29^ However, that explanation is probably unlikely for strongly influential species. *Proteus mirabilis*, *Staphylococcus aureus*, and *Anaerococcus hydrogenalis* were among the most abundant species and had the highest correlations with wound healing time among the parceled species. Each of these species has been shown to be drivers of infection of chronic wounds. *S. aureus* is the leading cause of delayed wound healing and has been shown to form biofilms, driving prolonged infection.^30^ *P. mirabilis* has also been shown to be a biofilm former and is the third main cause of hospital-acquired infections of immunocompromised patients.^31^ *A. hydrogenalis* has been previously reported in wounds and its inclusion here may partially reflect genus-level phenotypes, as *Anaerococcus* has been reported in roughly 50% of chronic deep tissue wounds.^32^ Because these bacteria are most associated with healing complications, it is initially paradoxical they were included in, and strongly influenced, the microbiome latent variable associated with faster time to heal. However, their inverse relationship with healing time (larger relative abundances at initial clinical visit were associated with shorter healing times) can be explained by the participating clinic’s adherence to biofilm-based wound care. Data for patients in this study are from their initial clinical visit, and patients are treated with topical compounds including biofilm dispersing agents in conjunction with multiple empiric or DNA sequencing-guided antibiotic therapies. Effectively treating infecting pathogens with this approach may explain why afore-mentioned influential species are inversely related to healing time, therefore including treatment as a latent variable will be explored in future studies utilizing this SEM framework.

The final model was validated by application to an independent cohort of 79 unique wounds at their first visit for treatment. Somewhat unexpectedly, the model had better predictive power on the validation cohort as compared to the initial cohort. This improvement is likely explained by reduced measurement error in data records for the validation cohort. The initial cohort of 565 unique wounds had the median of initial visits occurring in 2017 (1^st^ Qu. = 2015, 3^rd^ Qu. = 2020) and encompassed changes in personnel and record keeping strategy. The validation cohort spanned a shorter period (Median = 2021, 1^st^ Qu. = 2018, 3^rd^ Qu. = 2022), during which the attending physician promoted rigorous standardization of data entry and conducted all initial patient evaluations themselves.

The parceling approach to integrating microbiome composition into SEM is expected to be useful in a variety of scenarios analyzing highly dimensional ‘-omics’ data, and there is no expectation that its utility is limited to the scope of chronic wounds nor even microbiome analysis. With respect to wound infection specifically, one limitation of the current study is that it focused on wound environment and wound microbiome at the first clinical visit for each patient, but a longitudinal perspective could be taken to better account for changes as the wound heals. For example, previous work on chronic wounds has demonstrated they can be compositionally variable over time,^6,12^ and longitudinal perspectives are expected to further improve modeling. Also, whereas the dominant effect of microbiome on healing can reasonably be interpreted as a product of a microbe-centric treatment philosophy, expanding to include data from clinics that do not practice biofilm-based wound care and assessing model performance could evaluate this notion. Finally, future modeling could explore incorporating information about antibiotics and other treatments administered, as well as antimicrobial resistance detection. Other example variables that could be integrated include readings from a pulse oximeter or patient genotype.^33^ Although there will be future opportunities for model improvement, the current study provides a strong demonstration that clinical data can be integrated with SEM to predict healing outcomes.

## Data Availability

All data produced in the present study are available upon reasonable request to the authors.

## ACKNOWLEDGEMENTS

We thank MicroGenDX and CEO Rick Martin for their support of this work. We thank the staff at the Southwest Regional Wound Care Center for their commitment to excellence in wound care, scientific collaboration, and logistical support. This work was supported by NIH award R15GM141973.

The authors Jacob Ancira and Craig Tipton are employed by, and Caleb Phillips is a consultant to, MicroGenDx at the time of submission.

## Notes

### Competing Interest Statement

The authors Jacob Ancira, Khalid Omeir and Craig Tipton are employed by, and Caleb Phillips is a consultant to, MicroGenDx at the time of submission.

### Funding Statement

This study was supported by NIH award R15GM141973.

### Author Declarations

Texas Tech University Human Research Protection Program gave ethical approval for this work under IRB2016-1073.

