## Supplemental Table 1 for "A Structural Equation Model Predicts Chronic Wound Healing Time Using Patient Characteristics and Wound Microbiome Composition"

Supplementary Table 1: Patient demographics and wound characteristics by wound type. Cell values are either means with parenthetical standard deviations, or n with parenthetical percent of total n. The correlation or cooccurrence with the microbiome latent variable is shown depending on the defined relationship in the structural equation model. Values under Slough, Exudate, Percent Granulation, and Edema are the transformed scores according to the TIME-H comorbidity score defined in the methods section.

|  | Decubitus Ulcer,  N = 42 | Diabetic Foot Ulcer,  N = 105 | Atypical Wound,  N = 385 | Venous Leg Ulcer,  N = 33 | Correlation with Shorter Healing Time Microbiome Variable |
| --- | --- | --- | --- | --- | --- |
| Duration (Days) | 293 (297) | 464 (568) | 399 (535) | 271 (330) | -0.6343 |
| Volume |  |  |  |  | -0.1723 |
| Length (cm) | 0.85 (1.82) | 0.48 (0.80) | 0.56 (1.12) | 0.75 (1.52) |  |
| Width (cm) | 0.82 (1.66) | 0.41 (0.53) | 0.54 (1.12) | 0.74 (1.44) |  |
| Depth (cm) | 0.33 (0.72) | 0.19 (0.14) | 0.22 (0.30) | 0.22 (0.29) |  |
| Slough |  |  |  |  | -0.4931 |
| 0 | 26 (62%) | 78 (74%) | 286 (74%) | 23 (70%) |  |
| 1 | 12 (29%) | 26 (25%) | 87 (23%) | 10 (30%) |  |
| 2 | 4 (9.5%) | 1 (1.0%) | 12 (3.1%) | 0 (0%) |  |
| Exudate |  |  |  |  | -0.3334 |
| 0 | 17 (40%) | 58 (55%) | 199 (52%) | 20 (61%) |  |
| 1 | 16 (38%) | 31 (30%) | 134 (35%) | 7 (21%) |  |
| 2 | 9 (21%) | 16 (15%) | 52 (14%) | 6 (18%) |  |
| Percent Granulation |  |  |  |  | 0.0934 |
| 0 | 24 (57%) | 70 (67%) | 242 (63%) | 29 (88%) |  |
| 1 | 1 (2.4%) | 0 (0%) | 6 (1.6%) | 0 (0%) |  |
| 2 | 17 (40%) | 35 (33%) | 137 (36%) | 4 (12%) |  |
| Edema |  |  |  |  | -0.3659 |
| 0 | 24 (57%) | 78 (74%) | 240 (62%) | 22 (67%) |  |
| 1 | 12 (29%) | 25 (24%) | 125 (32%) | 11 (33%) |  |
| 2 | 6 (14%) | 2 (1.9%) | 20 (5.2%) | 0 (0%) |  |
