## Supplemental Table 2 for "A Structural Equation Model Predicts Chronic Wound Healing Time Using Patient Characteristics and Wound Microbiome Composition"

Supplementary Table 2: The table shows the number of wounds each of the species were found in. All wounds counted in the table were not polymicrobial. The table has also been facetted by the wound type.

| **Species** | **Overall,**  **N = 68** | **Decubitus Ulcer,**  **N = 3** | **Diabetic Foot Ulcer,**  **N = 11** | **Atypical Wound,**  **N = 49** | **Venous Leg Ulcer,**  **N = 5** |
| --- | --- | --- | --- | --- | --- |
| *Burkholderia cepacia* | 1 (1.5%) | 0 (0%) | 0 (0%) | 0 (0%) | 1 (20%) |
| *Corynebacterium jeikeium* | 1 (1.5%) | 0 (0%) | 1 (9.1%) | 0 (0%) | 0 (0%) |
| *Corynebacterium striatum* | 5 (7.4%) | 0 (0%) | 1 (9.1%) | 4 (8.2%) | 0 (0%) |
| *Enterococcus faecalis* | 1 (1.5%) | 0 (0%) | 0 (0%) | 1 (2.0%) | 0 (0%) |
| *Escherichia coli* | 1 (1.5%) | 0 (0%) | 0 (0%) | 1 (2.0%) | 0 (0%) |
| *Klebsiella aerogenes* | 1 (1.5%) | 0 (0%) | 0 (0%) | 1 (2.0%) | 0 (0%) |
| *Marinobacter hydrocarbonoclasticus* | 1 (1.5%) | 0 (0%) | 1 (9.1%) | 0 (0%) | 0 (0%) |
| *Pseudomonas aeruginosa* | 8 (12%) | 0 (0%) | 1 (9.1%) | 6 (12%) | 1 (20%) |
| *Staphylococcus aureus* | 32 (47%) | 3 (100%) | 7 (64%) | 19 (39%) | 3 (60%) |
| *Staphylococcus epidermidis* | 8 (12%) | 0 (0%) | 0 (0%) | 8 (16%) | 0 (0%) |
| *Staphylococcus lugdunensis* | 2 (2.9%) | 0 (0%) | 0 (0%) | 2 (4.1%) | 0 (0%) |
| *Staphylococcus pseudintermedius* | 1 (1.5%) | 0 (0%) | 0 (0%) | 1 (2.0%) | 0 (0%) |
| *Streptococcus agalactiae* | 4 (5.9%) | 0 (0%) | 0 (0%) | 4 (8.2%) | 0 (0%) |
| *Streptococcus anginosus* | 1 (1.5%) | 0 (0%) | 0 (0%) | 1 (2.0%) | 0 (0%) |
| *Streptococcus dysgalactiae* | 1 (1.5%) | 0 (0%) | 0 (0%) | 1 (2.0%) | 0 (0%) |
