## Supplemental Table 3 for "A Structural Equation Model Predicts Chronic Wound Healing Time Using Patient Characteristics and Wound Microbiome Composition"

Supplementary Table 3: List of the 66 parceled species incidence per wound type. Additionally, the species correlation with wound healing time and the respective parcel each species was included in is shown. The species pearson correlation with healing time was calculated during parceling procedure defined in the methods.

| **Species** | **Overall, N = 565** | **Decubitus Ulcer,**  **N = 42** | **Diabetic Foot Ulcer,**  **N = 105** | **Atypical Wound, N = 385** | **Venous Leg Ulcer,**  **N = 33** | **Healing Time Pearson Correlation** | **Parcel** |
| --- | --- | --- | --- | --- | --- | --- | --- |
| *Achromobacter xylosoxidans* | 3 (0.5%) | 0 (0%) | 1 (1.0%) | 2 (0.5%) | 0 (0%) | -0.0396 | p2 |
| *Acinetobacter lwoffii* | 4 (0.7%) | 2 (4.8%) | 0 (0%) | 2 (0.5%) | 0 (0%) | -0.0358 | p1 |
| *Acinetobacter radioresistens* | 4 (0.7%) | 0 (0%) | 0 (0%) | 3 (0.8%) | 1 (3.0%) | -0.0406 | p1 |
| *Actinobaculum massiliense* | 5 (0.9%) | 2 (4.8%) | 2 (1.9%) | 1 (0.3%) | 0 (0%) | -0.0234 | p2 |
| *Actinomyces neuii* | 14 (2.5%) | 1 (2.4%) | 5 (4.8%) | 7 (1.8%) | 1 (3.0%) | -0.0404 | p0 |
| *Actinotignum schaalii* | 3 (0.5%) | 0 (0%) | 1 (1.0%) | 2 (0.5%) | 0 (0%) | -0.0272 | p0 |
| *Alcaligenes faecalis* | 6 (1.1%) | 1 (2.4%) | 2 (1.9%) | 3 (0.8%) | 0 (0%) | -0.0271 | p2 |
| *Anaerococcus hydrogenalis* | 30 (5.3%) | 2 (4.8%) | 8 (7.6%) | 17 (4.4%) | 3 (9.1%) | -0.0634 | p0 |
| *Anaerococcus lactolyticus* | 38 (6.7%) | 7 (17%) | 7 (6.7%) | 23 (6.0%) | 1 (3.0%) | -0.0580 | p1 |
| *Anaerococcus octavius* | 10 (1.8%) | 0 (0%) | 0 (0%) | 10 (2.6%) | 0 (0%) | -0.0464 | p0 |
| *Anaerococcus prevotii* | 26 (4.6%) | 1 (2.4%) | 4 (3.8%) | 18 (4.7%) | 3 (9.1%) | -0.0541 | p1 |
| *Bacteroides dorei* | 2 (0.4%) | 0 (0%) | 0 (0%) | 2 (0.5%) | 0 (0%) | -0.0253 | p2 |
| *Bacteroides fragilis* | 13 (2.3%) | 1 (2.4%) | 0 (0%) | 12 (3.1%) | 0 (0%) | -0.0594 | p0 |
| *Bacteroides ovatus* | 2 (0.4%) | 1 (2.4%) | 0 (0%) | 1 (0.3%) | 0 (0%) | -0.0247 | p0 |
| *Bacteroides pyogenes* | 4 (0.7%) | 0 (0%) | 0 (0%) | 4 (1.0%) | 0 (0%) | -0.0205 | p2 |
| *Brevibacterium luteolum* | 3 (0.5%) | 0 (0%) | 0 (0%) | 3 (0.8%) | 0 (0%) | -0.0372 | p1 |
| *Brevundimonas nasdae* | 2 (0.4%) | 0 (0%) | 0 (0%) | 2 (0.5%) | 0 (0%) | -0.0259 | p1 |
| *Campylobacter ureolyticus* | 12 (2.1%) | 2 (4.8%) | 3 (2.9%) | 7 (1.8%) | 0 (0%) | -0.0470 | p2 |
| *Clostridium bolteae* | 2 (0.4%) | 1 (2.4%) | 0 (0%) | 1 (0.3%) | 0 (0%) | -0.0278 | p1 |
| *Corynebacterium afermentans* | 4 (0.7%) | 1 (2.4%) | 0 (0%) | 3 (0.8%) | 0 (0%) | -0.0276 | p1 |
| *Corynebacterium amycolatum* | 12 (2.1%) | 3 (7.1%) | 2 (1.9%) | 7 (1.8%) | 0 (0%) | -0.0288 | p0 |
| *Corynebacterium auriscanis* | 3 (0.5%) | 0 (0%) | 1 (1.0%) | 2 (0.5%) | 0 (0%) | -0.0398 | p0 |
| *Corynebacterium confusum* | 2 (0.4%) | 0 (0%) | 0 (0%) | 2 (0.5%) | 0 (0%) | -0.0239 | p2 |
| *Corynebacterium fournierii* | 2 (0.4%) | 0 (0%) | 0 (0%) | 2 (0.5%) | 0 (0%) | -0.0214 | p2 |
| *Corynebacterium pseudodiphtheriticum* | 4 (0.7%) | 1 (2.4%) | 1 (1.0%) | 2 (0.5%) | 0 (0%) | -0.0488 | p0 |
| *Corynebacterium striatum* | 60 (11%) | 1 (2.4%) | 15 (14%) | 38 (9.9%) | 6 (18%) | -0.0323 | p0 |
| *Corynebacterium tuscaniense* | 2 (0.4%) | 0 (0%) | 0 (0%) | 2 (0.5%) | 0 (0%) | -0.0288 | p2 |
| *Cutibacterium acnes* | 12 (2.1%) | 1 (2.4%) | 0 (0%) | 10 (2.6%) | 1 (3.0%) | -0.0554 | p0 |
| *Dialister micraerophilus* | 6 (1.1%) | 0 (0%) | 0 (0%) | 6 (1.6%) | 0 (0%) | -0.0334 | p0 |
| *Dialister pneumosintes* | 5 (0.9%) | 0 (0%) | 1 (1.0%) | 4 (1.0%) | 0 (0%) | -0.0512 | p0 |
| *Escherichia coli* | 60 (11%) | 6 (14%) | 7 (6.7%) | 43 (11%) | 4 (12%) | -0.0308 | p1 |
| *Faecalibacterium prausnitzii* | 2 (0.4%) | 0 (0%) | 1 (1.0%) | 1 (0.3%) | 0 (0%) | -0.0228 | p2 |
| *Fusobacterium canifelinum* | 5 (0.9%) | 0 (0%) | 3 (2.9%) | 2 (0.5%) | 0 (0%) | -0.0445 | p2 |
| *Fusobacterium nucleatum* | 26 (4.6%) | 1 (2.4%) | 7 (6.7%) | 18 (4.7%) | 0 (0%) | -0.0230 | p1 |
| *Fusobacterium periodonticum* | 6 (1.1%) | 0 (0%) | 2 (1.9%) | 4 (1.0%) | 0 (0%) | -0.0203 | p1 |
| *Gemella morbillorum* | 7 (1.2%) | 0 (0%) | 4 (3.8%) | 3 (0.8%) | 0 (0%) | -0.0303 | p1 |
| *Klebsiella aerogenes* | 2 (0.4%) | 0 (0%) | 0 (0%) | 2 (0.5%) | 0 (0%) | -0.0314 | p1 |
| *Klebsiella oxytoca* | 6 (1.1%) | 0 (0%) | 0 (0%) | 6 (1.6%) | 0 (0%) | -0.0298 | p0 |
| *Klebsiella pneumoniae* | 10 (1.8%) | 1 (2.4%) | 1 (1.0%) | 8 (2.1%) | 0 (0%) | -0.0310 | p2 |
| *Morganella morganii* | 15 (2.7%) | 3 (7.1%) | 3 (2.9%) | 8 (2.1%) | 1 (3.0%) | -0.0394 | p1 |
| *Pasteurella dagmatis* | 2 (0.4%) | 0 (0%) | 0 (0%) | 2 (0.5%) | 0 (0%) | -0.0283 | p0 |
| *Pasteurella multocida* | 6 (1.1%) | 1 (2.4%) | 1 (1.0%) | 4 (1.0%) | 0 (0%) | -0.0453 | p2 |
| *Peptoniphilus harei* | 35 (6.2%) | 5 (12%) | 6 (5.7%) | 23 (6.0%) | 1 (3.0%) | -0.0447 | p2 |
| *Peptostreptococcus anaerobius* | 16 (2.8%) | 1 (2.4%) | 2 (1.9%) | 11 (2.9%) | 2 (6.1%) | -0.0376 | p0 |
| *Porphyromonas bennonis* | 39 (6.9%) | 4 (9.5%) | 7 (6.7%) | 27 (7.0%) | 1 (3.0%) | -0.0329 | p2 |
| *Porphyromonas cangingivalis* | 4 (0.7%) | 0 (0%) | 1 (1.0%) | 3 (0.8%) | 0 (0%) | -0.0399 | p2 |
| *Porphyromonas levii* | 18 (3.2%) | 3 (7.1%) | 4 (3.8%) | 11 (2.9%) | 0 (0%) | -0.0445 | p1 |
| *Porphyromonas somerae* | 24 (4.2%) | 3 (7.1%) | 1 (1.0%) | 19 (4.9%) | 1 (3.0%) | -0.0513 | p1 |
| *Prevotella bergensis* | 9 (1.6%) | 2 (4.8%) | 0 (0%) | 7 (1.8%) | 0 (0%) | -0.0497 | p2 |
| *Prevotella corporis* | 6 (1.1%) | 2 (4.8%) | 1 (1.0%) | 3 (0.8%) | 0 (0%) | -0.0213 | p0 |
| *Prevotella timonensis* | 27 (4.8%) | 5 (12%) | 2 (1.9%) | 20 (5.2%) | 0 (0%) | -0.0205 | p1 |
| *Proteus mirabilis* | 28 (5.0%) | 2 (4.8%) | 5 (4.8%) | 19 (4.9%) | 2 (6.1%) | -0.1191 | p2 |
| *Proteus vulgaris* | 2 (0.4%) | 0 (0%) | 0 (0%) | 1 (0.3%) | 1 (3.0%) | -0.0238 | p2 |
| *Staphylococcus arlettae* | 2 (0.4%) | 0 (0%) | 0 (0%) | 2 (0.5%) | 0 (0%) | -0.0244 | p2 |
| *Staphylococcus aureus* | 195 (35%) | 14 (33%) | 35 (33%) | 133 (35%) | 13 (39%) | -0.1057 | p1 |
| *Staphylococcus capitis* | 16 (2.8%) | 1 (2.4%) | 3 (2.9%) | 11 (2.9%) | 1 (3.0%) | -0.0473 | p0 |
| *Staphylococcus caprae* | 3 (0.5%) | 0 (0%) | 2 (1.9%) | 1 (0.3%) | 0 (0%) | -0.0208 | p0 |
| *Staphylococcus cohnii* | 9 (1.6%) | 1 (2.4%) | 3 (2.9%) | 5 (1.3%) | 0 (0%) | -0.0468 | p1 |
| *Staphylococcus pseudintermedius* | 4 (0.7%) | 0 (0%) | 2 (1.9%) | 2 (0.5%) | 0 (0%) | -0.0424 | p0 |
| *Stenotrophomonas maltophilia* | 9 (1.6%) | 0 (0%) | 2 (1.9%) | 6 (1.6%) | 1 (3.0%) | -0.0365 | p1 |
| *Streptococcus dysgalactiae* | 18 (3.2%) | 1 (2.4%) | 2 (1.9%) | 14 (3.6%) | 1 (3.0%) | -0.0396 | p1 |
| *Streptococcus intermedius* | 6 (1.1%) | 1 (2.4%) | 0 (0%) | 5 (1.3%) | 0 (0%) | -0.0569 | p2 |
| *Streptococcus mitis* | 17 (3.0%) | 2 (4.8%) | 4 (3.8%) | 11 (2.9%) | 0 (0%) | -0.0394 | p2 |
| *Streptococcus oralis* | 6 (1.1%) | 0 (0%) | 0 (0%) | 5 (1.3%) | 1 (3.0%) | -0.0418 | p0 |
| *Streptococcus pyogenes* | 4 (0.7%) | 0 (0%) | 3 (2.9%) | 1 (0.3%) | 0 (0%) | -0.0214 | p1 |
| *Terrahaemophilus aromaticivorans* | 5 (0.9%) | 0 (0%) | 1 (1.0%) | 4 (1.0%) | 0 (0%) | -0.0343 | p0 |
